# Renin-angiotensin-aldosterone system inhibitors and COVID-19 infection or hospitalization: a cohort study

**DOI:** 10.1101/2020.07.06.20120386

**Authors:** Sascha Dublin, Rod Walker, James S. Floyd, Susan M. Shortreed, Sharon Fuller, Ladia Albertson-Junkans, Laura B. Harrington, Mikael Anne Greenwood-Hickman, Beverly B. Green, Bruce M. Psaty

## Abstract

There are plausible mechanisms by which angiotensin-converting enzyme inhibitors (ACEIs) and angiotensin receptor blockers (ARBs) may increase the risk of COVID-19 infection or affect disease severity. To examine the association between these medications and COVID-19 infection or hospitalization, we conducted a retrospective cohort study within a US integrated healthcare system. Among people aged ≥18 years enrolled in the health plan for at least 4 months as of 2/29/2020, current ACEI and ARB use was identified from pharmacy data, and the estimated daily dose was calculated and standardized across medications. COVID-19 infections were identified through 6/14/2020 from laboratory and hospitalization data. We used logistic regression to estimate adjusted odds ratios (ORs) and 95% confidence intervals. Among 322,044 individuals, 720 developed COVID-19 infection. Among people using ACEI/ARBs, 183/56,105 developed COVID-19 (3.3 per 1000 individuals) compared with 537/265,939 without ACEI/ARB use (2.0 per 1000), yielding an adjusted OR of 0.94 (95% CI 0.75-1.16). For use of < 1 defined daily dose vs. nonuse, the adjusted OR for infection was 0.89 (95% CI 0.62-1.26); for 1 to < 2 defined daily doses, 0.97 (95% CI 0.71-1.31); and for ≥2 defined daily doses, 0.94 (95% CI 0.72-1.23). The OR was similar for ACEIs and ARBs and in subgroups by age and sex. 29% of people with COVID-19 infection were hospitalized; the adjusted OR for hospitalization in relation to ACEI/ARB use was 0.92 (95% CI 0.54-1.57), and there was no association with dose. These findings support current recommendations that individuals on these medications continue their use.

## Introduction

Use of angiotensin converting enzyme inhibitors (ACEIs) and angiotensin receptor blockers (ARBs), prescribed for nearly 25% of US adults,^1^ may be a risk factor for coronavirus disease 2019 (COVID-19) because these drugs increase the expression of angiotensin converting enzyme 2 (ACE2),^2^ the receptor by which the SARS-CoV-2 coronavirus enters epithelial cells.^3^ Concern about whether inhibitors of the renin-angiotensin-aldosterone system (RAAS) may increase susceptibility to COVID-19 has been so pronounced that professional societies have issued advisories urging patients not to discontinue them and calling for more evidence.^4^

On the other hand, experimental evidence suggests that upregulation of ACE2 may protect against lung injury caused by severe coronavirus infection.^5^ Among hospitalized patients with COVID-19 and hypertension, those on ACEI/ARBs had lower levels of high-sensitivity C-reactive protein and procalcitonin.^6^ Most observational studies have focused on people hospitalized for COVID-19, examining whether ACEI/ARB use is associated with worse clinical outcomes.^6-9^ While this sampling design addresses important questions, it selects for patients who are already infected and whose disease has become severe enough to require hospitalization, Such studies cannot shed light on the natural history of infection prior to hospitalization or whether the use of RAAS inhibitors may increase susceptibility to COVID-19. Three studies have examined the risk of infection in relation to RAAS use among people from well-characterized populations with information about prior medication exposures and health conditions.^10-12^ These studies, set in Italy,^10^ Spain^11^ and Denmark,^12^ found no overall association between RAAS inhibitor use and COVID-19 infection. Testing patterns and case fatality rates may vary widely between countries.^13^ No true population-based study has yet been conducted in the US, which has a very different health care system and more racially diverse population than these European countries. While rigorous, these studies lacked information about smoking status, obesity and race/ethnicity, which may be important confounders,^14-16^ and their COVID cases were disproportionately weighted towards hospitalized cases – lacking cases from the milder end of the spectrum. Finally, no study has yet examined the relationship between ACEI/ARB dose and risk of COVID-19 infection or severe disease.

In a population-based setting with rich electronic health resources, we evaluated the associations of ACEI and ARB use including medication dose with the risk of COVID-19 infection and, as a marker of severity, with hospitalization.

## Methods

We conducted a retrospective cohort study within Kaiser Permanente Washington (KPWA), an integrated health care system in Washington State that maintains extensive electronic data on its members. Members within the integrated group practice (IGP) receive all or nearly all care from KPWA. When they need hospitalization, members are cared for at contracted hospitals. Because reimbursement depends on these records, data about these hospitalizations tend to be very accurate. The population eligible for these analyses was IGP members who were aged ≥ 18 years and enrolled in KPWA in February 2020 (the month before COVID-19 testing began at KPWA). To be included, members had to have at least 4 months of prior enrollment as of 2/29/2020 and additional enrollment beyond that date. The sample size was determined by the number of eligible. Individuals were followed for study outcomes through 6/14/2020. Study procedures were approved by the KPWHRI Institutional Review Board with a waiver of consent.

We used electronic pharmacy data to define exposure to RAAS inhibitors. Pharmacy data come from KPWA-owned pharmacies and in addition include medications dispensed at outside pharmacies (via claims data). Data were available in near-real-time, including dispensings up through the day prior to the data pull. Data include a member identification number, date of the fill, medication name and strength, number of pills dispensed, and estimated days’ supply. “Current use” was defined as having a dispensed medication with a supply sufficient to last until 2/29/2020 or later, assuming 80% adherence. To estimate the daily dose, we multiplied the number of pills dispensed by pill strength and divided by the estimated days’ supply for the dispensing. We then standardized these daily doses across medications by dividing by the World Health Organization’s Defined Daily Dose for each medication (e.g., 10 mg of lisinopril).^17^

We defined COVID-19 infection as having a positive COVID-19 reverse-transcriptase polymerase chain reaction (PCR) test or a hospitalization with a COVID-19 diagnosis code (patients admitted to contracted hospitals for severe illness would not necessarily have had prior COVID-19 testing at KPWA). Throughout the study period, KPWA followed Washington State testing guidelines. For much of this time, the guidelines recommended testing only individuals with severe illness and symptomatic patients with high-risk exposures or health conditions.^18^ To assess whether these apparent COVID-19 hospitalizations represented true cases, we reviewed medical records for 20 patients hospitalized with a COVID-19 diagnosis who did not have a positive COVID-19 PCR test within KPWA.

We identified covariates from KPWA electronic health data. Demographic characteristics included age, sex, and self-reported race/ethnicity. We ascertained chronic medical conditions using diagnosis codes from inpatient and ambulatory visits; a condition was considered to be present if an individual had one or more codes for that condition in any setting in the prior 12 months. Use of other medications was determined from electronic pharmacy data. The KPWA EHR includes information about tobacco use (which patients are routinely asked about at clinic visits) and body mass index (BMI), calculated from clinical measures of height and weight. Medications of interest included prednisone, insulin, and other classes of antihypertensive medications (calcium channel blockers, beta blockers, thiazide diuretics, and loop diuretics.) See the Supplement for detailed variable specifications.

Within the entire cohort, we used logistic regression to estimate the odds of COVID-19 infection associated with RAAS inhibitor use with adjustment for age (<45, 45-64, 65+ years), sex, race/ethnicity (categorized as non-Hispanic white, non-Hispanic Black, non-Hispanic Asian, non-Hispanic mixed or other race, or Hispanic), diabetes, hypertension, heart failure (HF), prior myocardial infarction (MI), asthma, chronic obstructive pulmonary disease (COPD), current tobacco use, renal disease, malignancy, Charlson comorbidity score^19^ (categorized as 0, 1, 2+), BMI (categorized as underweight, <18.5 kg/m^2^; normal weight, 18.5-24.9 kg/m^2^; overweight, 25-29.9 kg/m^2^; obese, 30-34.9 kg/m^2^; and severely obese, ≥ 35 kg/m^2^), and use of insulin, loop diuretics, and prednisone. Among those with COVID-19 infection, we used logistic regression to estimate the odds of hospitalization associated with RAAS inhibitor use with adjustment for a reduced set of covariates. To account for missing data on race/ethnicity (13%) and BMI (27%), we used multiple imputation by chained equations^20^ to generate 25 datasets with missing information imputed. Logistic regression models were estimated using these datasets and results were pooled following Rubin’s rules.^21^

We conducted analyses examining the association between ACEI/ARB daily dose and risk of infection or hospitalization. The referent category was no use of ACEI/ARBs. The standardized daily dose was grouped as < 1, 1 to <2, and ≥ 2 defined daily doses (DDDs) per day.

Secondary analyses for both infection and hospitalization risk evaluated associations for ACEI and ARB use separately and in subgroups by sex and age (< 65 or ≥ 65 years). For the latter, we included interaction terms between each factor and RAAS inhibitor use in the models and tested significance with a Wald test of the interaction term. In sensitivity analyses, we examined other antihypertensive medications as “control exposures” to evaluate whether associations were specific to RAAS inhibitors. Because many chronic health conditions are indications for both COVID-19 testing and RAAS inhibitor use, we repeated our primary analyses of COVID-19 infection limited to individuals who underwent testing and separately, to those with an indication for RAAS inhibitors (hypertension, diabetes, HF or prior MI). For risk of infection, we conducted additional sensitivity analyses restricted to individuals taking at least one antihypertensive medication, adjusted for the number of medications.

Analyses were conducted using R version 3.5.3 (Vienna, Austria) including the mice package (version 3.8.0).

## Results

Figure 1 shows the selection of the study sample. There were 322,044 eligible individuals; their mean age was 51 years, 74% were non-Hispanic white and 46% were male. 56,105 (17%) were current users of ACEIs or ARBs. Lisinopril accounted for 96% of ACEI fills and losartan 97% of ARB fills. Individuals using ACEI/ARBs were older than nonusers, more likely to be obese, and more likely to have many chronic conditions, consistent with indications for RAAS inhibitor use (Table 1). Among ACEI/ARB users, 21% had a low daily dose (< 1 DDD per day), 32% had a medium dose (1 to < 2 DDD/day) and 47% a high dose (≥ 2 DDD per day).

**Table 1.**
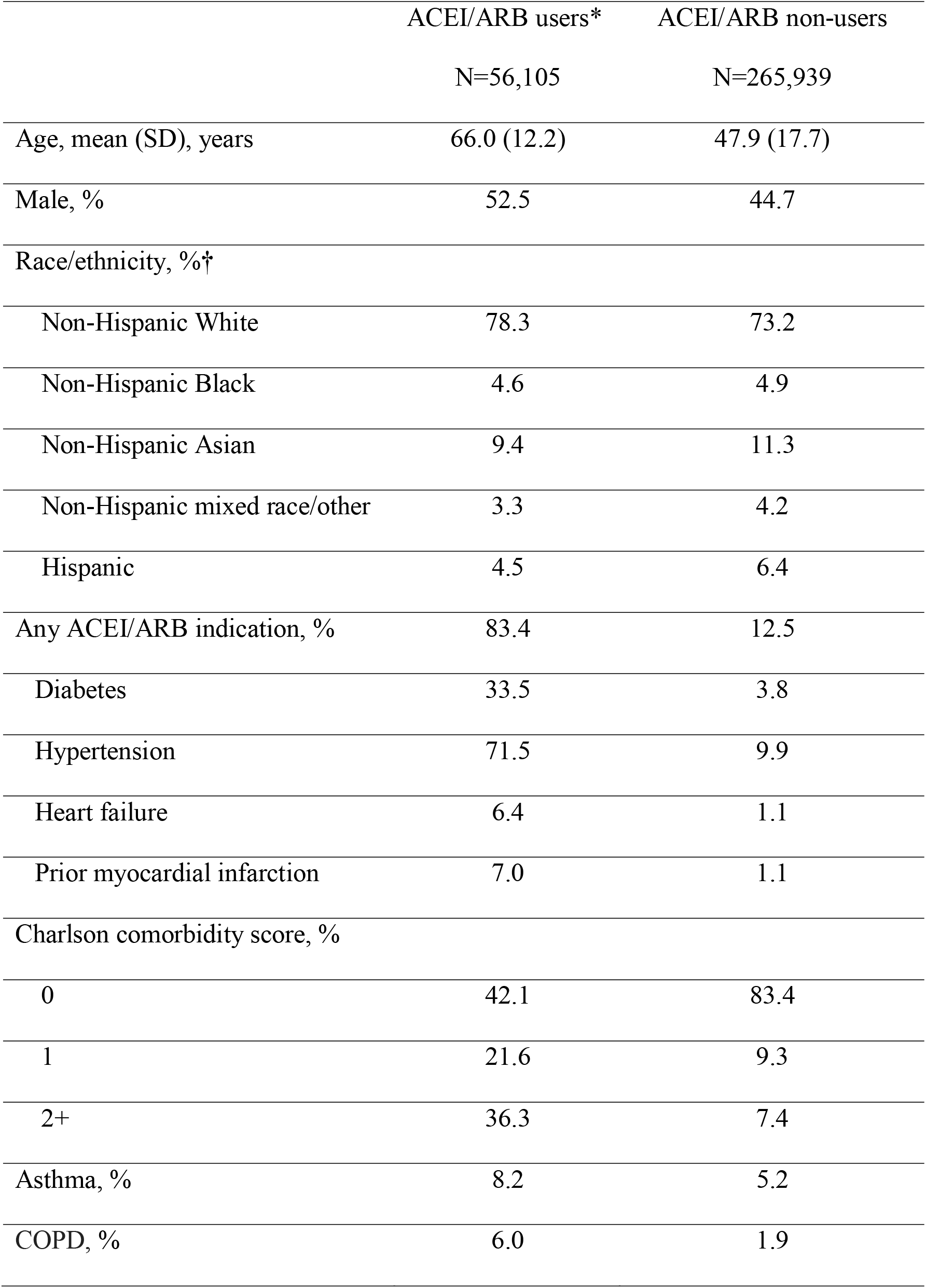
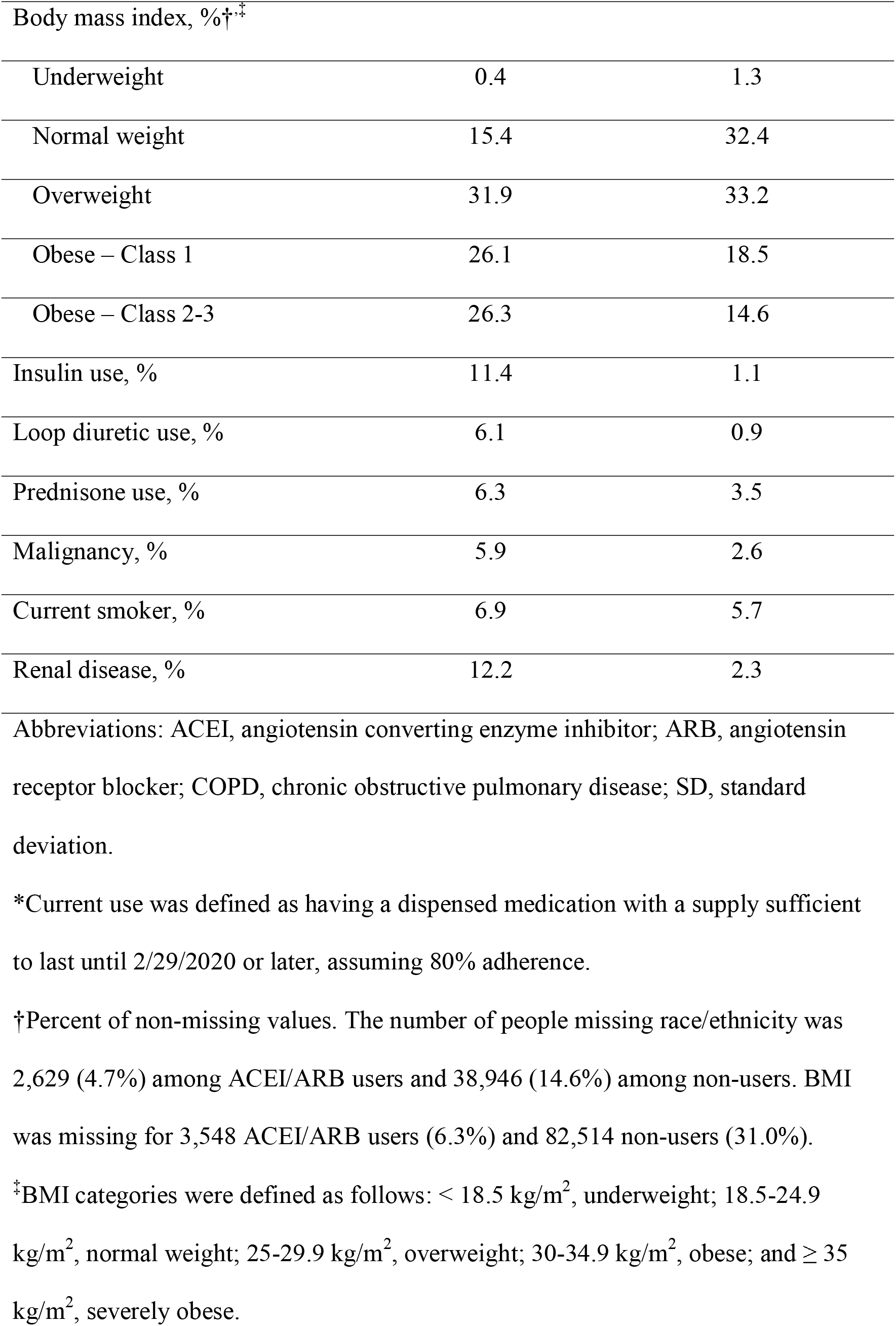
Characteristics of study population by ACEI/ARB use.

**Figure 1.**
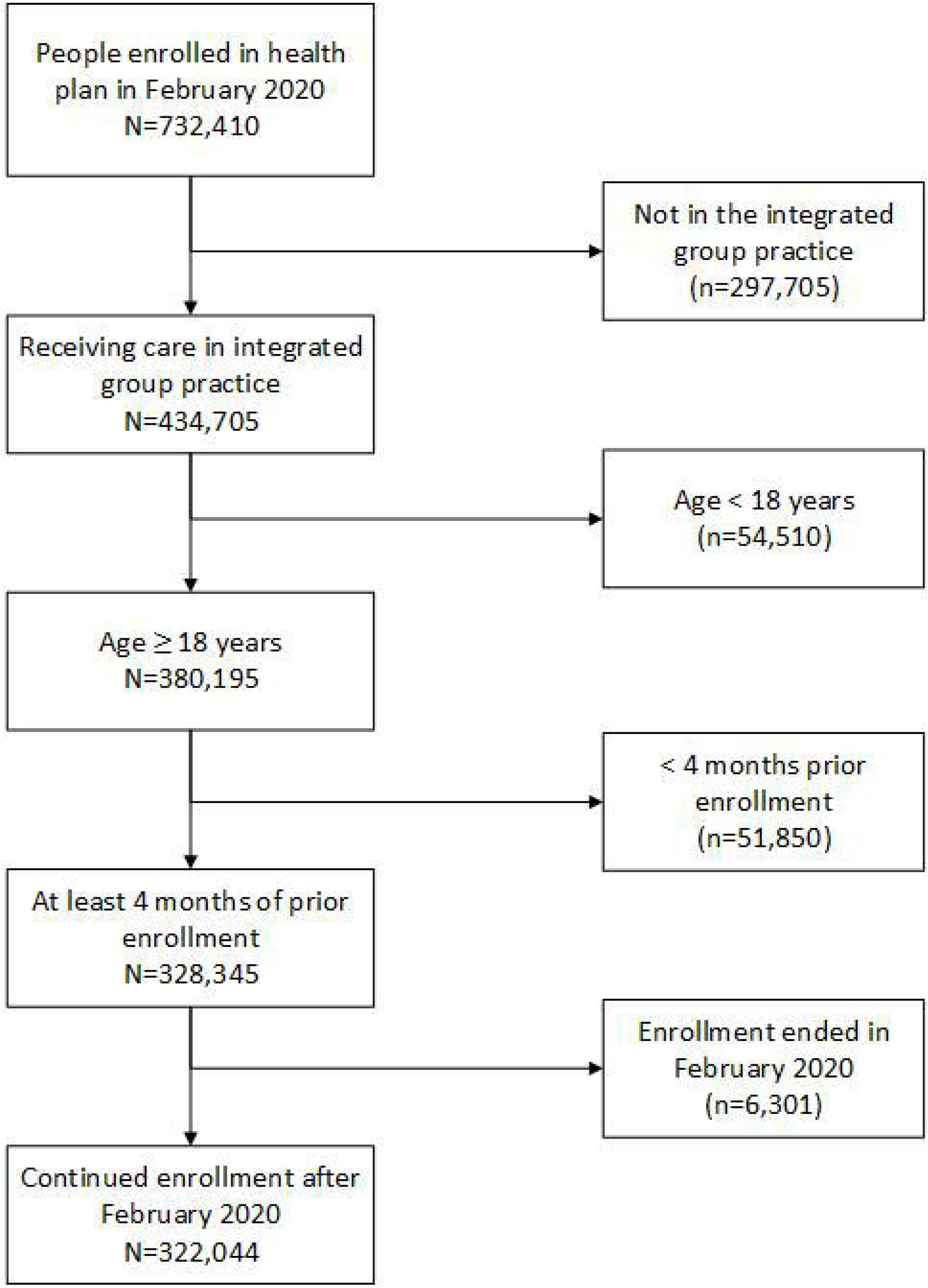
Selection of the study cohort.

Among 14,547 people tested for COVID-19 between 2/29/2020 and 6/14/2020, 597 (4.1%) tested positive. An additional 123 individuals were hospitalized with a diagnosis of COVID-19, for a total of 720 COVID-19 infections. For a sample of 20 COVID-19 hospitalizations without a positive test at KPWA, we reviewed medical records and confirmed positive PCR results at outside institutions for 17 (85%).

Among individuals with RAAS inhibitor use, 183/56,105 developed COVID-19 infection (3.3 per 1000 individuals) compared with 537/265,939 among nonusers (2.0 per 1000). The unadjusted OR for COVID-19 infection in relation to RAAS inhibitor use was 1.62 (95% CI 1.37-1.91). After adjustment for covariates, the OR was 0.94 (95% CI 0.75-1.16). Risk of infection was strongly associated with race/ethnicity and obesity (Table 2). There was no association between the dose of ACEI/ARB and risk of infection (Figure 2); the adjusted OR for people taking a low dose (< 1 DDD per day) compared to nonusers was 0.89 (95% CI 0.62-1.26), for a medium dose (1 to < 2 DDD) it was 0.97 (95% CI 0.71-1.31), and for a high dose (≥ 2 DDD), 0.94 (95% CI 0.72-1.23).

**Table 2.**
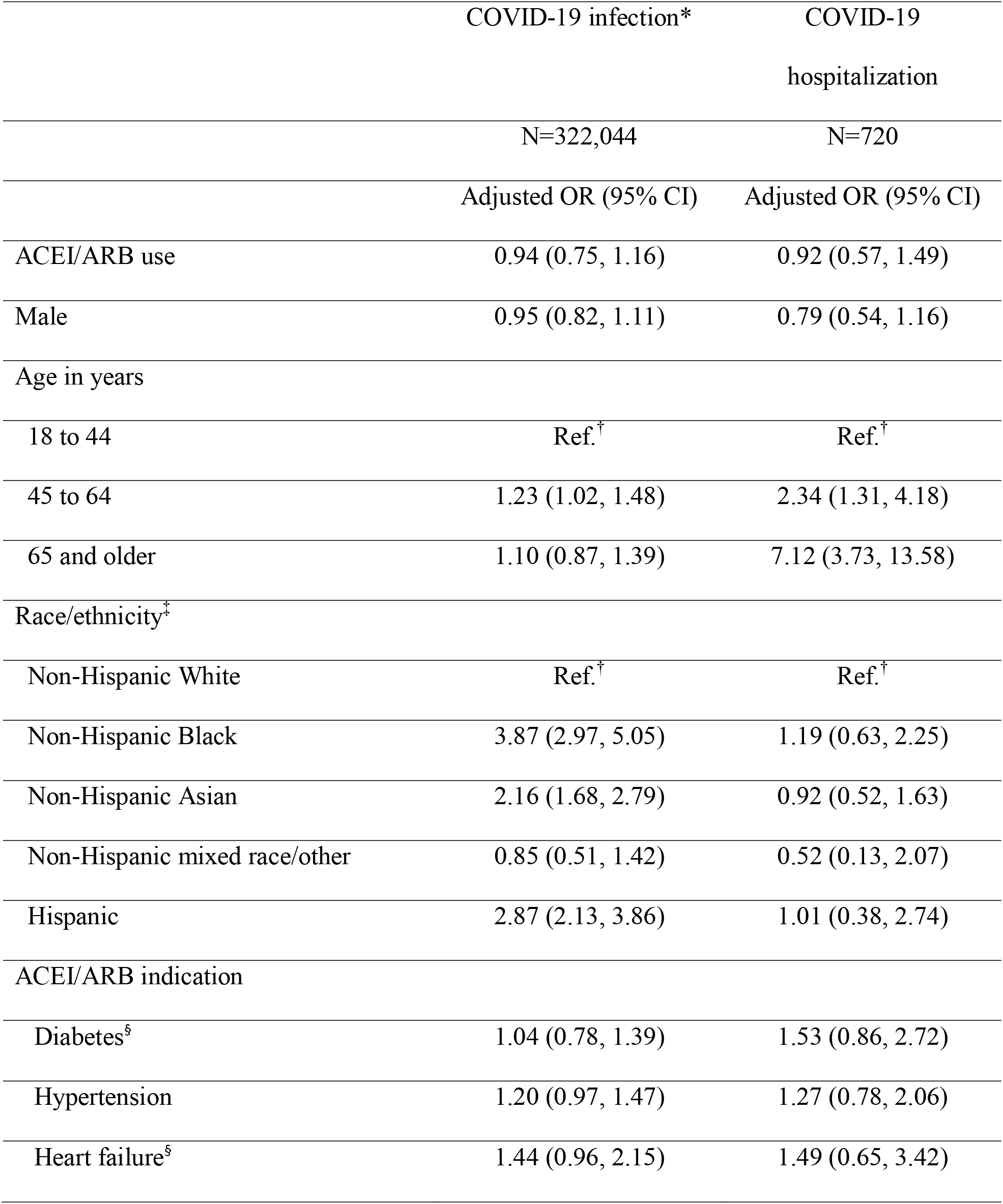
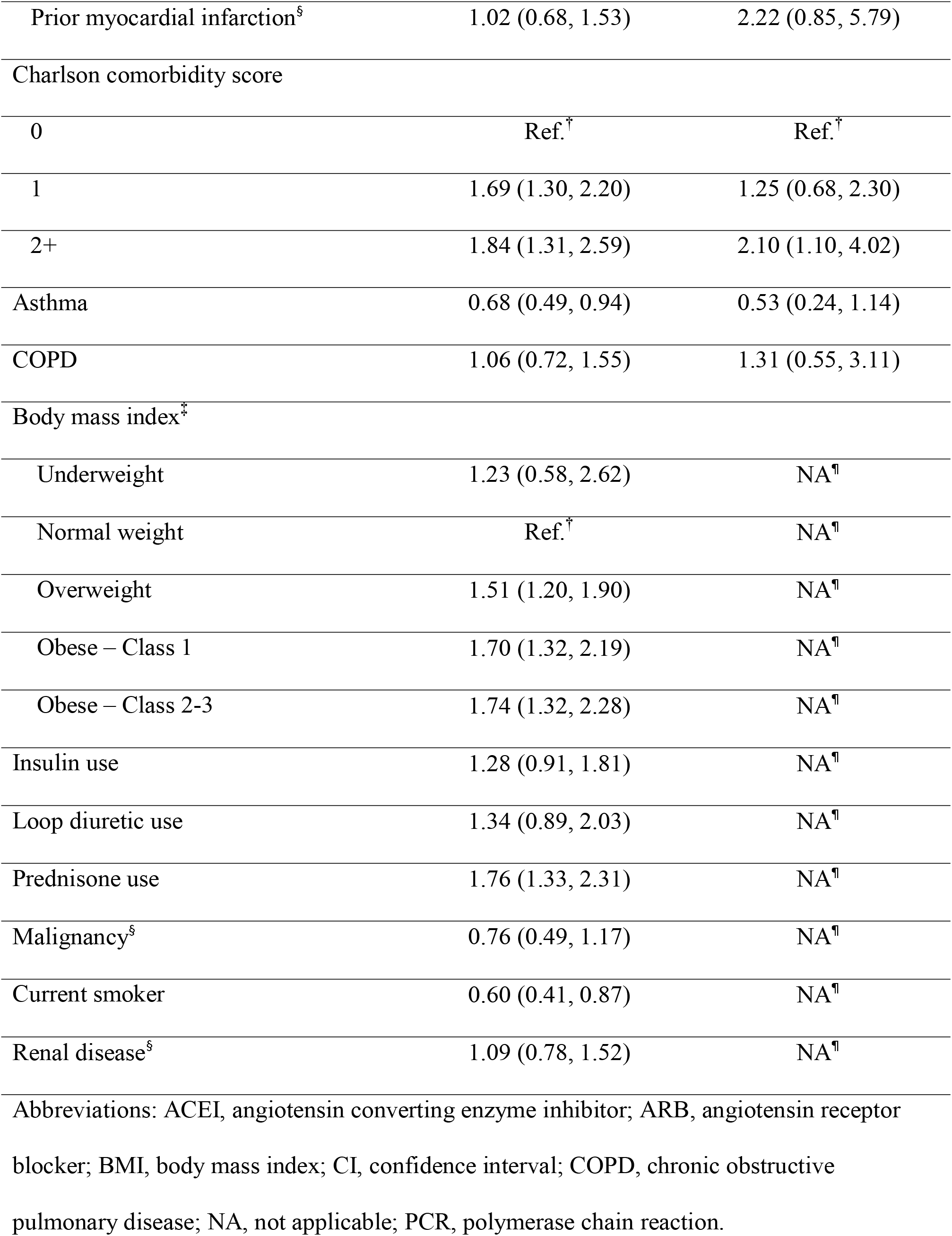
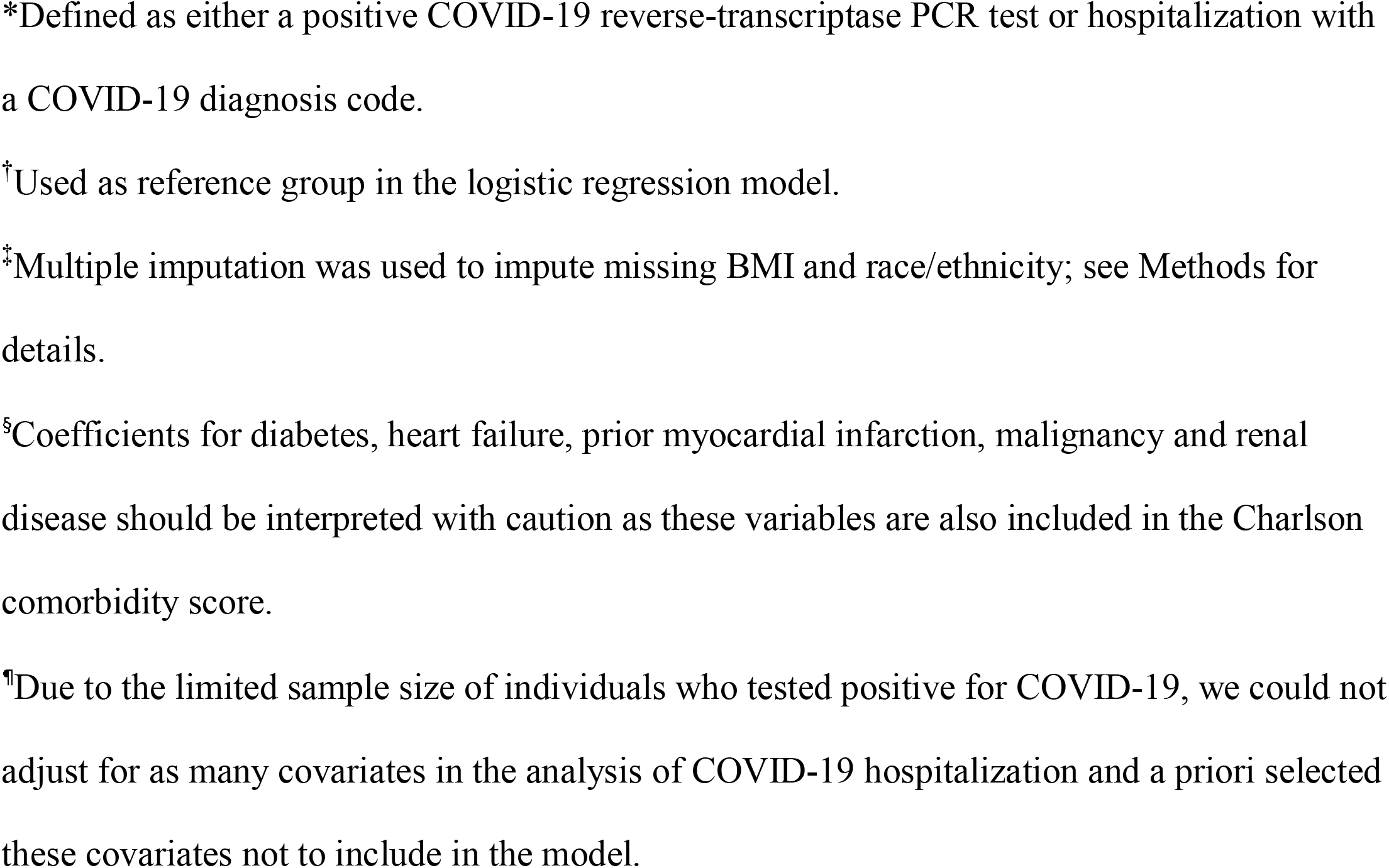
Associations of ACEI/ARB use with risk of COVID-19 infection and hospitalization.

**Figure 2.**
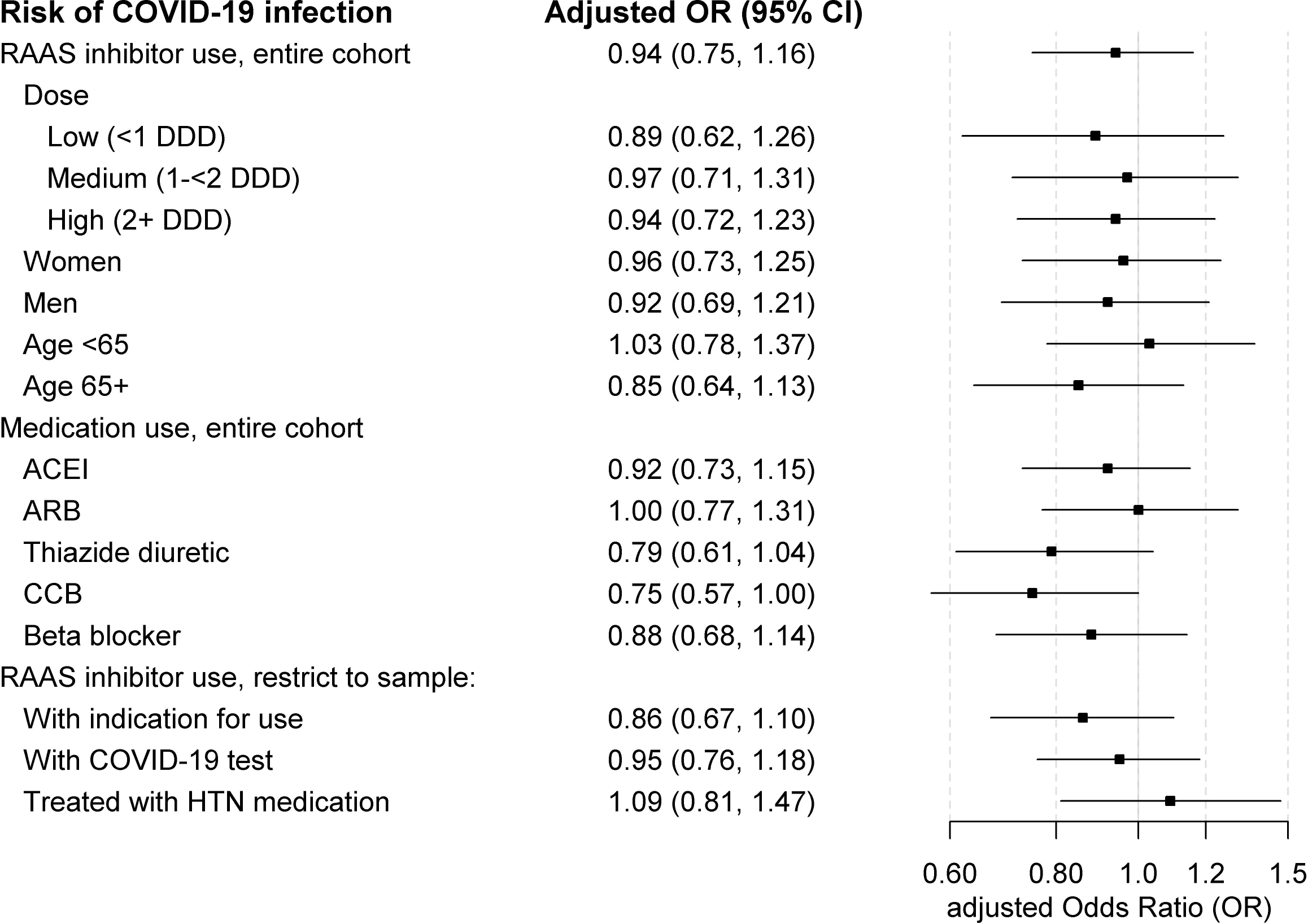
Odds of COVID-19 infection in relation to use of RAAS inhibitors. Estimates are adjusted for age, sex, race/ethnicity, diabetes, hypertension, HF, prior MI, asthma, COPD, current tobacco use, renal disease, malignancy, Charlson comorbidity score, BMI, and use of insulin, loop diuretics, and prednisone. Abbreviations: ACEI, angiotensin converting enzyme inhibitor; aOR, adjusted odds ratio; ARB, angiotensin receptor blocker; CCB, calcium channel blocker; CI, confidence interval; HTN, hypertension; RAAS, renin-angiotensin-aldosterone system.

Among individuals with COVID-19 infection, 211/720 (29.3%) were hospitalized, including 83/183 (45.4%) among RAAS inhibitor users and 128/537 (23.8%) among nonusers. The unadjusted OR for hospitalization comparing ACEI/ARB use to nonuse was 2.65 (95% CI 1.86-3.77), and the fully adjusted OR was 0.92 (95% CI 0.57-1.49). No association was seen between ACEI/ARB dose and hospitalization (Figure 3); for people taking a high daily dose, the adjusted OR for hospitalization was 0.86 (95% CI 0.47-1.57) compared to nonuse.

**Figure 3.**
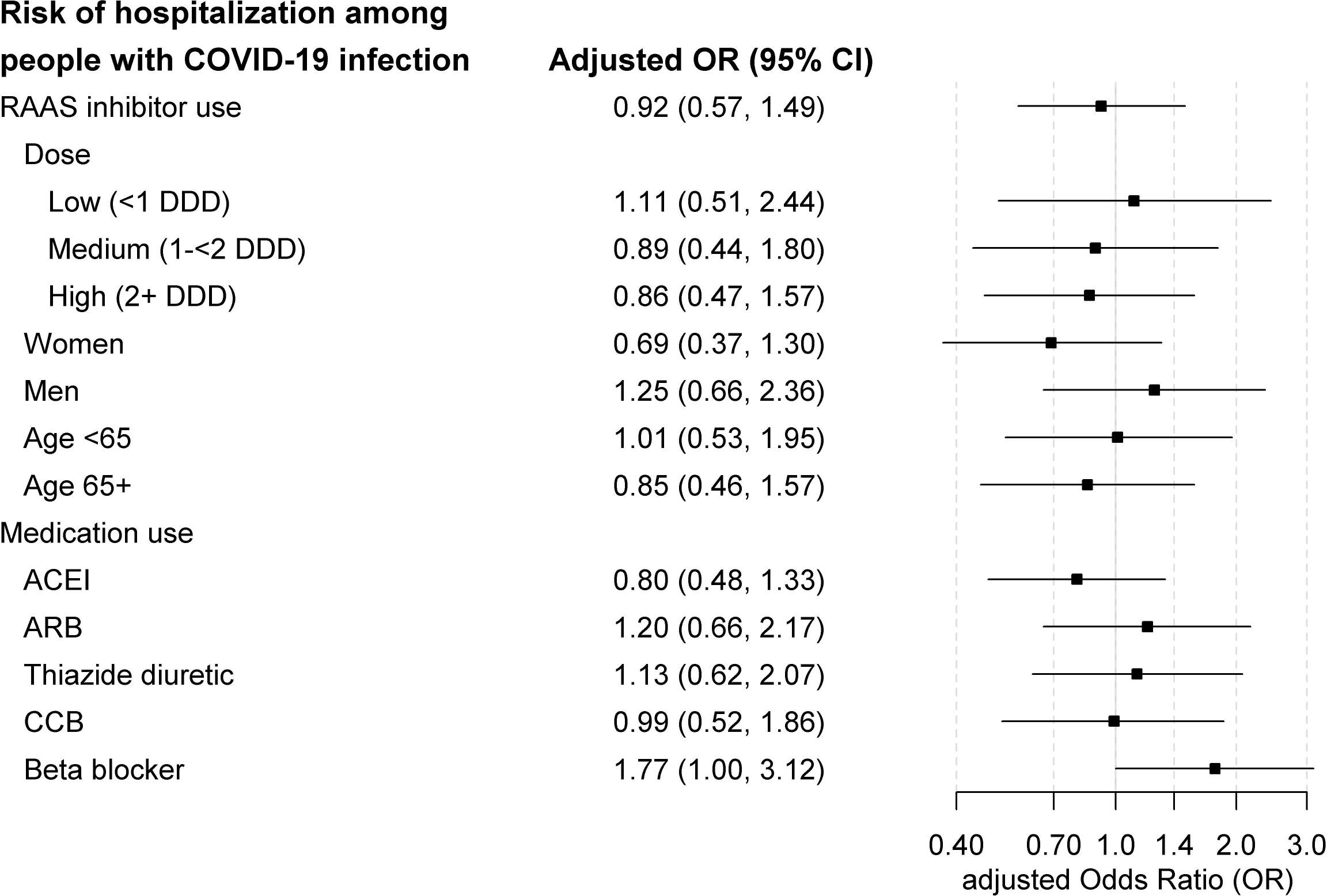
Odds of COVID-19 hospitalization in relation to use of RAAS inhibitors, among individuals with COVID-19 infection. Estimates are adjusted for age, sex, race/ethnicity, diabetes, hypertension, HF, prior MI, asthma or COPD, and Charlson comorbidity score. Abbreviations: ACEI, angiotensin converting enzyme inhibitor; aOR, adjusted odds ratio; ARB, angiotensin receptor blocker; CCB, calcium channel blocker; CI, confidence interval; RAAS, renin-angiotensin-aldosterone system.

In sensitivity analyses, the adjusted OR for COVID-19 infection was similar for ACEI and ARB users and in subgroups by age and sex (Figure 2). Risk estimates for ACEIs and ARBs were slightly higher than for thiazides, beta-blockers or calcium channel blockers (Figure 2). Findings changed little after restricting to individuals with an indication for RAAS inhibitor therapy, those tested for COVID-19, or those treated with antihypertensive medications. In sensitivity analyses examining COVID-19 hospitalization (Figure 3), results were similar for ACEIs and ARBs and in subgroups by age. Results appeared modestly different by sex, with an adjusted OR for ACEI/ARB use that was lower in women than in men, but this difference was not statistically significant (p = 0.16).

## Conclusions

In this population-based cohort study set within a US healthcare system, there was no significant association between use of RAAS inhibitors and the risk of COVID-19 infection or hospitalization, including no association of these outcomes with ACEI/ARB daily dose. This is the first study to our knowledge that has examined the association of medication dose with COVID-19 outcomes.

Most published studies have focused on the risk of complications among hospitalized patients.^6-9^ Our finding for infection risk is consistent with several other population-based studies,^10-12^ including a case-control study from Italy where the adjusted OR for infection in relation to ACEI/ARB use was 0.95 (95% CI 0.86-1.05)^10^ and a study from Denmark where the adjusted OR was 1.05 (95% CI 0.80-1.36).^12^ Examining this question in the US is important because of differences in the clinical context, COVID-19 testing practices and case fatality rates,^13^ and the distribution of race/ethnicity in the population.

A recent systematic review assessed the association between RAAS inhibitor use and COVID-19 infection and outcomes;^22^ they concluded that there is “moderate-certainty evidence” from 3 studies that ACEI/ARB use does not increase infection risk and stronger evidence that ACEI/ARB use does not increase the risk of severe outcomes. They noted limitations of prior studies including that many were small or did not adjust for important confounders, and that for some of the smaller studies, it was unclear how prior RAAS inhibitor use was ascertained. The 3 large population-based studies^10-12^ of this question (one of which was included in the systematic review^10^) were all set in Europe. They have many strengths and offer important evidence, but gaps still remain. When identifying COVID-19 infections, one study focused solely on hospitalized cases^11^ and a second ascertained only patients diagnosed in hospitals or hospital-based clinics.^12^ The cases identified thus represent more severe cases on the disease spectrum, and there remains a need for studies including cases with milder disease diagnosed in the ambulatory setting. The definition of exposure varied across studies, with potential for misclassification, and none of these studies included information about ACEI or ARB daily dose.

This study provides additional evidence that RAAS inhibitors do not increase the risk of SARS-CoV-2 infection. Strengths include the ability to examine the risk of COVID-19 infection including both outpatient and hospitalized cases arising in a well-defined population with extensive EHR data and complete capture of clinical events. We were able to estimate people’s daily dose of ACEI/ARBs and to adjust for characteristics typically missing from large healthcare databases such as smoking, BMI and race/ethnicity, characteristics which seem to have strong associations with risk of COVID-19 infection or more severe disease.^14-16^ Longitudinal data available for patients for years prior to the start of follow up improve the accuracy of ascertaining chronic illnesses and prior medication use. Limitations of observational studies such as this one include measurement error, selection bias, and residual confounding. Because KPWA testing practices followed state guidelines, we lack information about milder cases or those in low-risk populations. We could be missing hospitalizations for people who tested positive during follow-up and were hospitalized after the end of our follow-up period.

### Perspectives

In conclusion, these findings do not support an adverse effect of RAAS inhibitors on the risk of COVID-19 infection or hospitalization, including among individuals taking the highest doses of these medications. Our results provide additional support for clinical guidelines recommending that patients currently taking these medications need not stop them.

## Data Availability

Upon request, the authors will freely share the study protocol and statistical code. After the primary paper has been published, study data could be shared with approved individuals for specific research purposes through written agreements with the authors and with Kaiser Permanente Washington Health Research Institute.

## Data Sharing

Upon request, the authors will freely share the study protocol and statistical code. Study data could be shared with approved individuals for specific research purposes through written agreements with the authors and with Kaiser Permanente Washington Health Research Institute.

## Novelty and Significance

### What is New?

- In a general population, people using angiotensin converting enzyme inhibitors and angiotensin receptor blockers (widely used blood pressure medications) were not at higher risk of getting COVID-19. People on the highest doses were not at higher risk.
- If they became infected, their risk of hospitalization was similar to that of people not on these medications, and this association did not differ by ACEI/ARB dose.

### What is Relevant?

- People with hypertension commonly use these medication classes.
- Because these medications may increase expression of ACE2, a protein through which COVID-19 enters cells, there was concern they might increase the risk of infection or severe outcomes. If so, people on the highest doses might be at the greatest risk.

### Summary

People taking angiotensin converting enzyme inhibitors and angiotensin receptor blockers, including those using high doses, can continue to take them without concern about higher risk of COVID 19 infection.

